# Prevalence of and risk factors for high-risk HPV and cervical cytological abnormalities in women living with HIV in Bali, Indonesia

**DOI:** 10.1101/2024.09.21.24314131

**Authors:** I Ketut Agus Somia, Made Lady Adelaida Purwanta, Ni Wayan Winarti, Ida Bagus Nyoman Putra Dwija, Desak Made Putri Pidari, A.A.S. Sawitri, Anak Agung Ayu Yuli Gayatri, I Nyoman Gede Budiana, Komang Januartha Putra Pinatih, Ketut Tuti Parwati Merati

## Abstract

**Background:** Women living with HIV face a higher risk of developing cervical cancer compared to those without HIV. However, comprehensive cervical cancer screening programs for this population are still lacking in Indonesia. This has resulted in many cases of late-stage cervical cancer being diagnosed, especially in Bali, which has experienced an increase in cases of HIV and cervical cancer. This study aimed to determine the prevalence of and risk factors for cervical cytological abnormalities in women living with HIV in Bali and to explore their relationship with high-risk HPV (HR-HPV) types.

**Methods:** This is a cross-sectional study with eligible participants recruited from outpatient HIV clinics in Bali. Between July to December 2023, participants were interviewed to collect demographic and historical medical information, followed by physical examination including collection of cervical swabs and blood samples. Pap smear sampling and swab collection using ThinPrep for cytology. HPV DNA was then identified by PCR and genotyped for HR-HPV 16,18,31,33,35,39,45,51,52,56,58,59,66,68. Blood samples were analyzed for CD4 and CD8 cell counts.

**Results:** A total of 245 women with HIV at median age of 38 years old (24-50 years) and with median time of ARV therapy of 7 years (0-18 years) were enrolled. Only 239 participants were included in the analysis for their valid initial results. Overall, 26 (10.87%) of samples showed abnormal cytology including 6 (2.5%) ASC-H, 9 (3.8%) ASC-US, 4 (1.7%) H-SIL and 7 (2.9%) L-SIL. Of the 58 (24%) that tested positive for HPV DNA, 18 (31%) samples had abnormal cytology. HPV 18 was the most common genotype detected (n=16 or 28%). Bivariate analysis revealed a significant association between positive HPV DNA and abnormal cytology, with those testing HPV-positive having seven times higher risk of ASC-US or greater (PR=7.022;95%CI=3.223-15.295). Multivariate regression identified having HPV 18 infection as an independent risk factor for abnormal cytology (ExpB=9.029;p=0.007), and a history of Pap smear screening associated with reduced risk of HR-HPV infection (ExpB=0.358;p=0.013).

**Conclusion:** In our study, 10.87% of women living with HIV had abnormal cytology and 24% had positive HPV DNA tests. HPV 18 was associated with a greater risk of abnormal cytology compared to other high-risk HPV strains, but our sample size was small. History of pap smear was also shown to decrease the risk of HR-HPV infection. The results underscore the need for increased vaccination of younger women and screening of all women living with HIV in Indonesia in order to improve their cervical health outcomes.

## Introduction

Cervical cancer is one of the most frequently detected cancers in women living with HIV and is classified as an AIDS-defining illness. From the meta-analysis of 24 studies shows that women living with HIV have six times higher risk of developing cervical cancer compared to those without HIV.(1) Globally, an estimated 6% of new cervical cancer cases in 2018 were diagnosed among women living with HIV and 5% of all cases were attributable to HIV infection.^1^ IARC 2021 data shows that in Indonesia, 0.3% of cervical cancer cases were caused by co-infection with HIV. Due to being immunocompromised, women living with HIV are more prone to high-risk HPV strains that often cause cervical cancer, this could even result in the persistence of the HPV infection.(2,3) This leads to the urgency in early detection and management of cervical cancer in the population of women living with HIV.

Cervical cancer screening in HIV population is crucial to avoid further complication and mortality. Screening is the main strategy to reduce cervical cancer incidence, by detecting pre-cancerous lesions that can be treated before evolving to cancer. This, which is followed by adequate management is one of two prevention tools recommended by WHO that are highly effective. Based on CDC guidelines, for woman with newly diagnosed HIV-infection, the first cervical cancer screening is recommended by the time of HIV diagnosis and repeated on 6 or 12 months after diagnosis.(4) Effectiveness of the disease’s management is influenced by not only the accuracy of the screening method but also by the coverage of the cervical cancer treatment. However, in reality many countries have yet conducted established screening program for cervical cancer in this specific population, especially those of developing countries.

Indonesia was still considered at a low rate of cervical cancer screening (12%) per 2020, resulting in an emergency situation for cervical cancer until recently.(5) Many patients came to the hospital with their end-stage cancer due to the late diagnosis. On the other hand, there has also been increasing number of HIV cases. Based on the recapitulation data of the early detection of cervical cancer in Indonesia 2016, the number of suspected cervical cancer in several provinces such as Jakarta were 269 cases, Bali with 254 cases and Bangka Belitung with 227 cases. The data showed that Bali was one of the areas with high cervical cancer rates in 2016. Furthermore, this number increased in 2019 for Bali with the incidence of cervical cancer of 437 cases. On the other hand, Bali was on the sixth position for the highest HIV cases in Indonesia with 28,376 cases until June 2022 as stated by the Indonesian Ministry of Health June 2022.(6)However, there has yet been any data on the number of cervical cancer in this HIV population.

A number of risk factors could also influence the risk of cervical cytological abnormalities and cervical cancer in women living with HIV. Besides late screening, it was thought that late ART initiation or at lower CD4 cell counts could increase the risk of developing cervical cancer. ART are expected to decrease viral load and help improve CD4 cell counts, thus enhancing clearance of HPV and reducing the incidence of cervical cytological abnormalities.(2) Therefore, inequities in access to effective cervical cancer screening and treatment of precancerous cervical lesions are likely to be the main driver of high invasive cervical cancer rates in women living with HIV.

Due to the reasoning above, we would like to determine the prevalence and risk factors of cervical cytological abnormalities in women living with HIV in Bali, Indonesia, as well as to find its association with the high-risk HPV possible of cervical cancer. This study will provide not only the actual data of the local population to define future action in eradicating cervical cancer, but also directly provide benefit in the form of early detection for women living with HIV in Bali.

## Methods

### Study design and setting

This research was an observational study with cross-sectional design carried out at Yayasan Kerti Praja HIV foundation Denpasar Bali clinic 26^th^ of July 2023 – 11^th^ of December 2023. The respondents were women living with HIV who had regular appointments at the HIV clinics in Bali. All participants gave their consent by signing on a written informed consent. This study has been granted an ethical statement from the Ethic Committee of Faculty of Medicine, Udayana University with the number: 1661/UN14.2.2.VII.14/LT/2023.

### Patient selection and sampling

We included all women living with HIV of age 18-50 years old that attended the clinics. The inclusion criteria: 1) women living with HIV of age 18-50 years old; 2) Indonesian nationality; 3) has had sexual intercourse and willing to give their informed consent and exclusion criteria: 1) mentally unstable; 2) cervical cancer patients; 3) on menstruation during data collection; 4) pregnant women; 5) history of previous pap smear and/or HPV testing in the last 6 months; 6) on treatment of all types of pre-malignancy or malignancy and/or had hysterectomy.

Number of samples were calculated based on the sample size formula for cross-sectional design of proportional outcome. With the level of acceptable error of 5% and the expected proportion in population of 19.05%,(7) and at the 5% types I error rate, the minimum number of samples required was 237. Potential candidates were approached with consecutive sampling.

We screened 300 eligible candidates of women living with HIV, where these women had no certain symptoms, but rather concern regarding their predisposition to the disease. The screening procedure was done in a similar way to one of the algorithms (Algorithm 3) in the WHO Guidelines of Cervical Cancer Screening.(8) For research purpose, all participants in the study went through cytology screening with ThinPrep pap smear and HPV DNA testing simultaneously. This was done altogether to give a complete picture of all participants involved in this study as part of the Indonesian ethnicity.

### Study procedures

Blood samples were analyzed for CD4 and CD8 cell counts using the flow cytometry method using FACSVia (R656874100022) / FASClyric tool (R659180000700)

### Cytology identification

Cytological tests were performed by liquid based preparation technique (ThinPrep, Hologic, Inc., USA). All specimens were taken using ThinPrep swab kit, processed with a ThinPrep machine, stained with Papaniculaou stain, and assessed microscopically by a pathologist based on The Bethesda System criteria.

### PCR for HPV DNA and genotyping

Vaginal swab samples were collected and inserted into ThinPrep cytology media (ThinPrep PreservCyt Solution1 REF 70097-005—Hologic, Inc. Marlborough, MA, USA). Subsequently, the fluid in the cytology tube was vortexed, and 1 ml of ThinPrep fluid was transferred to a microcentrifuge tube. Centrifugation was performed at 12,000 RPM for 1 minute, and the pellet was washed with distilled water. DNA extraction was carried out using the Zymo Quick-DNATM Miniprep Plus Kit according to the manufacturer’s instructions. The extracted DNA was then stored at -20°C until use. Next, PCR was performed with a total volume of 10 µl, consisting of 10 nM forward and reverse primers, PCR mix, and distilled water. PCR was conducted in two stages; the first stage used the MY09/11 universal primers, and positive samples were followed by type-specific PCR using specific primers for HPV 16,18,31,33,35,39,45,51,52,58,59,66,68, and 69. PCR was carried out on a VeritiTM machine (AB Biosystem) with a denaturation PCR program of 95°C for 5 minutes, followed by 35 cycles of 95°C for 1 minute, 95°C for 20 seconds, 55°C for 45 seconds, 72°C for 60 seconds, and a final extension at 72°C for 7 minutes. PCR results were visualized using 1.5% agarose gel with GelRed staining under UV light

### Statistics

Initially, descriptive statistics were used to summarize the demographic and clinical characteristics of the study population. The prevalence of cervical abnormalities and HPV infection rates were calculated. Bivariate analysis was conducted to explore the association between HPV DNA positivity and abnormal cytology results. The strength of this association was quantified using odd ratio (ORs) with corresponding 95% confidence intervals (CIs). Multivariate logistic regression analysis was then performed to identify independent risk factors for cervical abnormalities, adjusting for potential confounders that had association in bivariate analysis of less than 0.25. The significance of multivariate analysis was determined with a threshold of p-value less than 0.05. We used SPSS version 29 to conduct the analysis.

## Results

From the targeted population, we received a list of 300 potential candidates for screening. After referred to the inclusion and exclusion criteria, we recruited 245 eligible participants, from which 239 samples (97.6%) were successfully analyzed. We had to repeat sampling 10 cases due to initial sampling inadequacy, attributed to challenging conditions such as difficult portio access or the presence of inflammatory cells. The research team gave significant efforts to communicate with participants requiring re-sampling, providing detailed explanations about the importance of the repeat pap smear procedure and the potential health benefits. Out of these ten needed to repeat, there were six samples excluded from the final analysis, two of these were due to suspected cervical cancer, and the other four were due to logistical issues (two participants relocated and could not return, and two declined to participate further despite multiple contact attempts, resulting in lost to follow up).The team still provided results from the initial tests along with health education and preventive advice concerning cervical cancer (see Figure 1).

**Figure 1.**
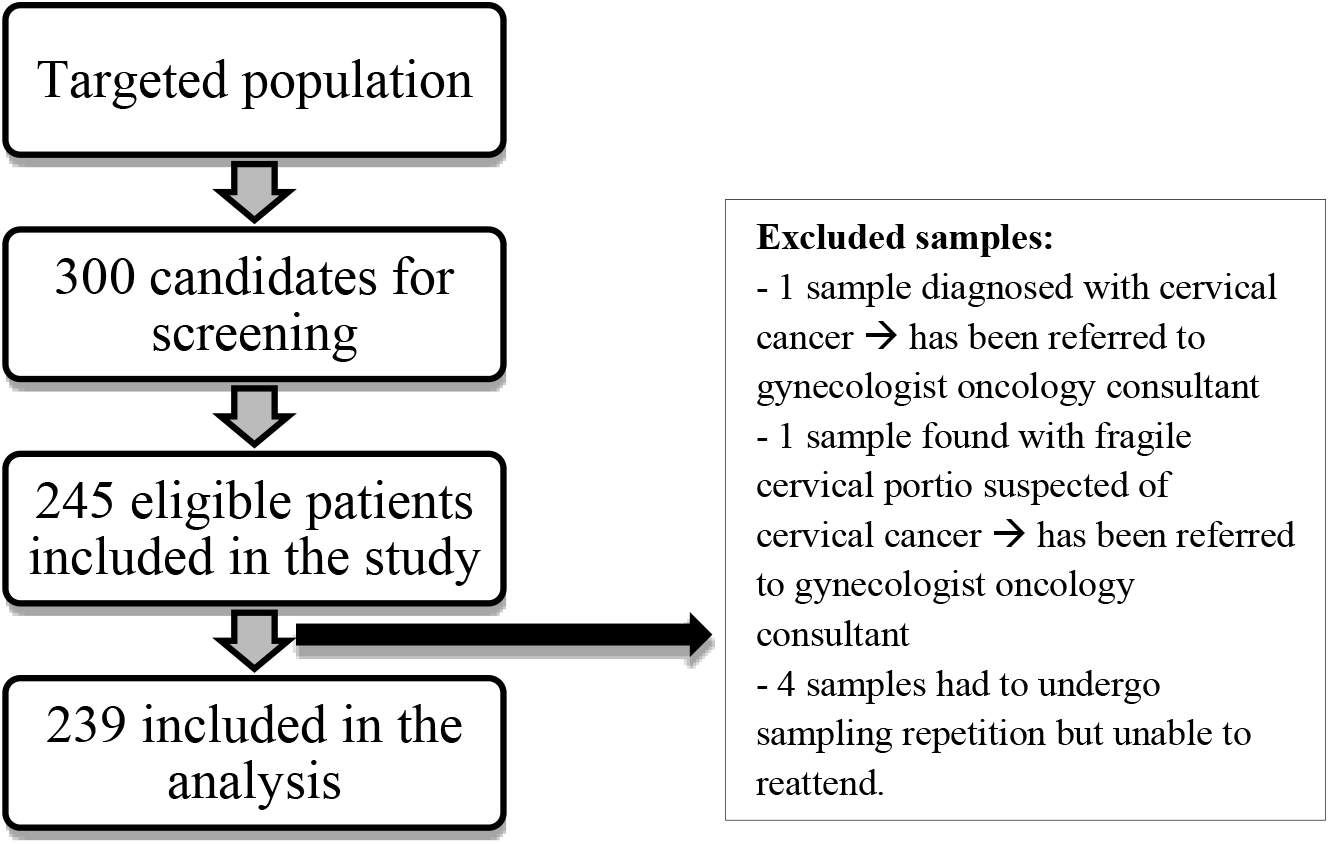
Recruitment process during the study.

### Baseline characteristics of study participants

Based on Table 1, the characteristics of study participants were grouped based on the presence or absence of cervical cytological abnormalities, categorized across various demographic and behavioral factors. The findings indicated that divorced individuals represented 8.8% of those with cytological abnormalities, while 91.3% did not have these conditions. Education levels revealed a slightly higher incidence cytological abnormalities among those with higher education, with those having completed high school or more showing a greater percentage compared to those with less education. Employment status correlated with cytological abnormalities prevalence, where 15.5% of unemployed participants had cytological abnormalities compared to 8.9% of employed participants. The analysis also considered lifestyle factors such as alcohol consumption and contraception use, noting that none of the alcohol consumers in the study had cytological abnormalities, and those using contraception exhibited a higher percentage of cytological abnormalities than those who did not. Sexual behavior was also a factor, with participants who had more sexual partners or an earlier sexual debut showing slightly higher percentages of cytological abnormalities. Additional factors like gravidity, hormonal medication use, and history of abortion were examined, each contributing to the understanding of how these characteristics were associated with the presence of ASC or cervical cytological abnormalities among the participants.

**Table 1.**
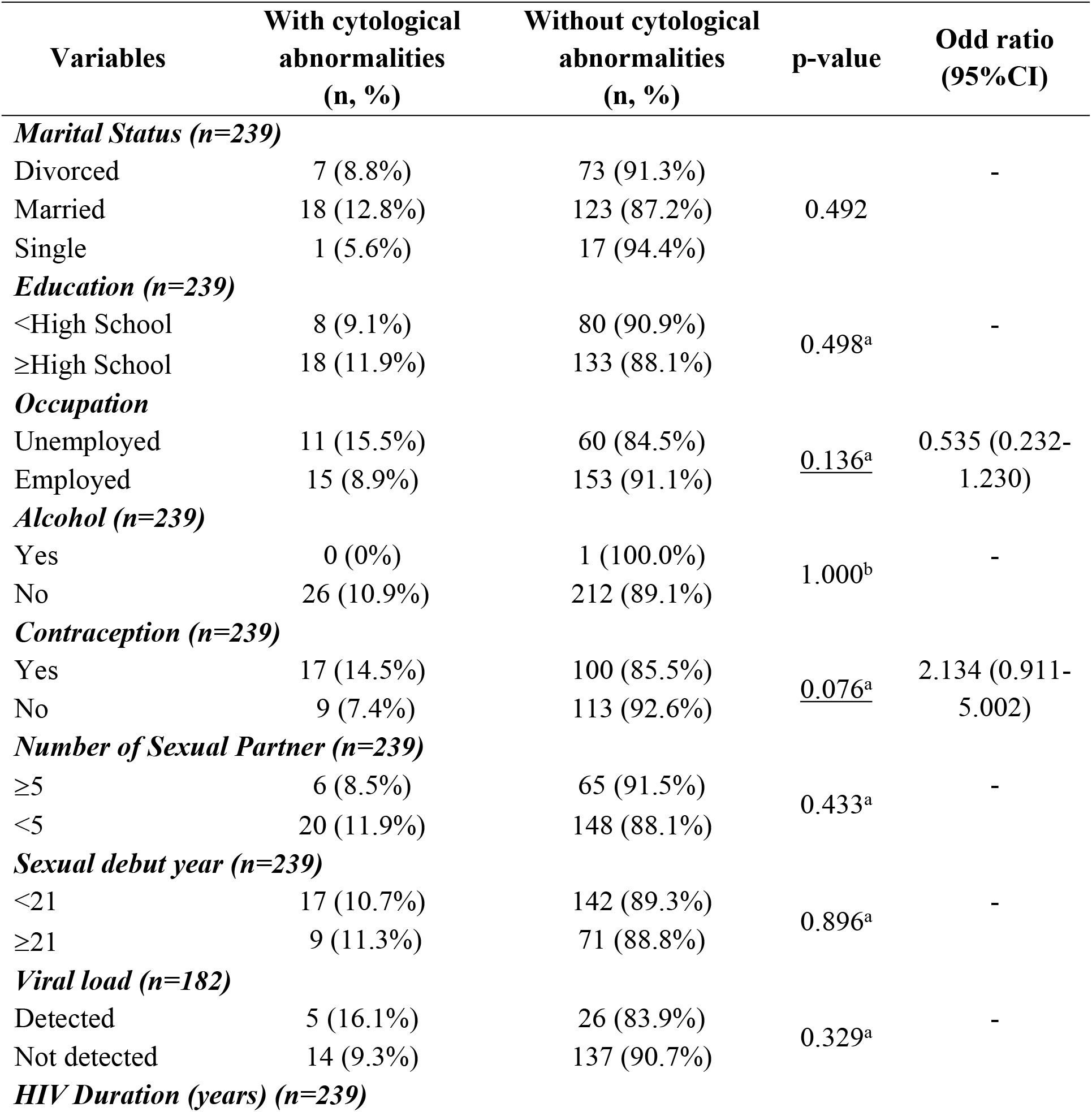
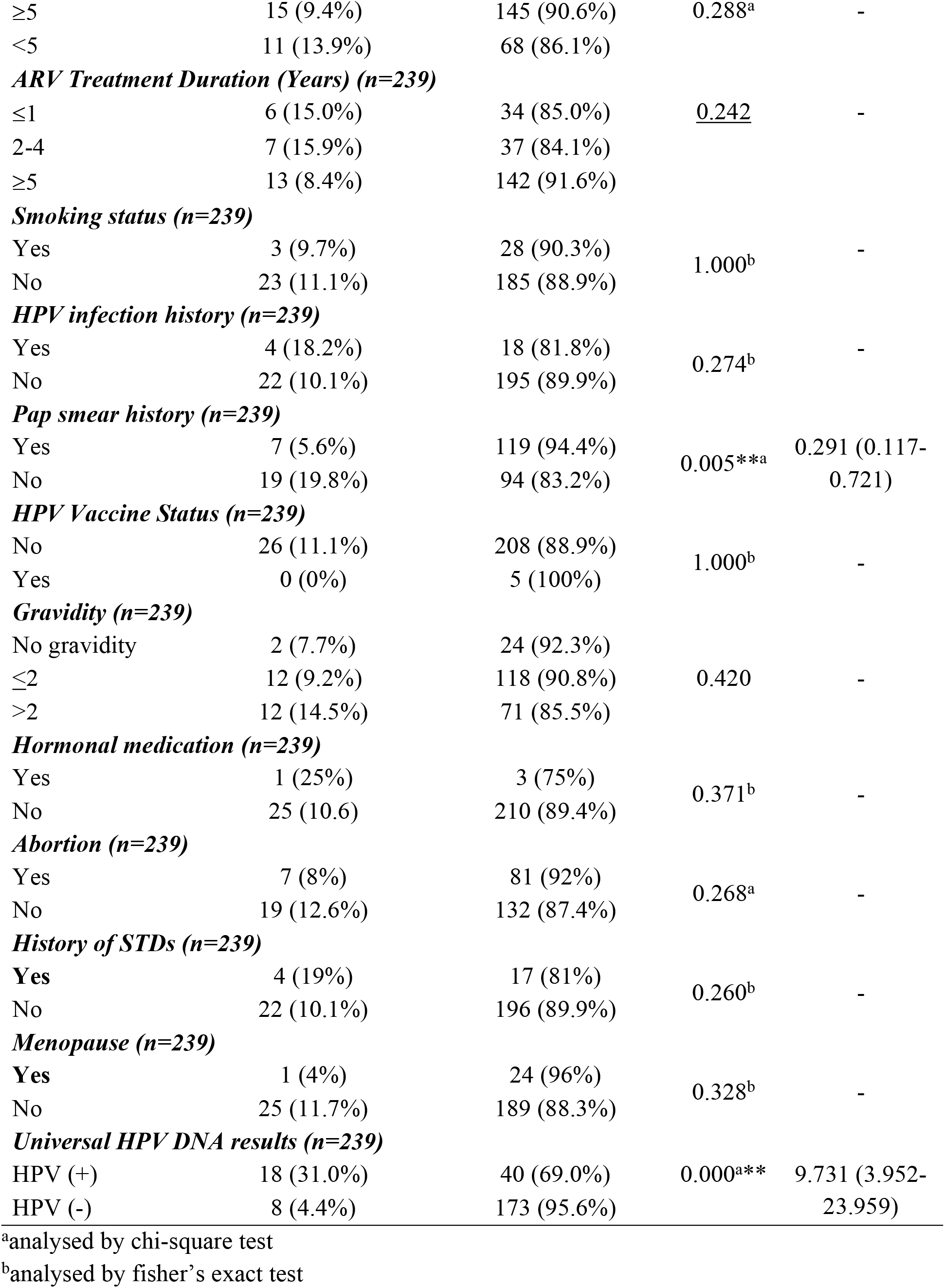
Characteristics of samples based on with and without cervical cytological abnormalities (categorical variables)

Based on Table 2, it was found that the average age of participants with cytological abnormalities was slightly younger at 35.5 years compared to 38 years for those without these conditions, although this difference was not statistically significant. Notably, the CD4 cell count showed a significant difference, with a median count of 415 cells/uL in those with Cytological abnormalities versus 556 cells/uL in those without, suggesting a lower immune status in the affected group. Similarly, the percentage of CD4 cells was significantly lower in participants with Cytological abnormalities, averaging 21.34%, compared to 26.62% in those without, further indicating compromised immune function in the former group. The CD8 cell count and percentage did not show significant differences. However, the CD4/CD8 ratio was significantly lower in participants with Cytological abnormalities, with a median of 0.43 compared to 0.63 in those without, underscoring a more pronounced immune system imbalance in those with cervical abnormalities.

**Table 2.**
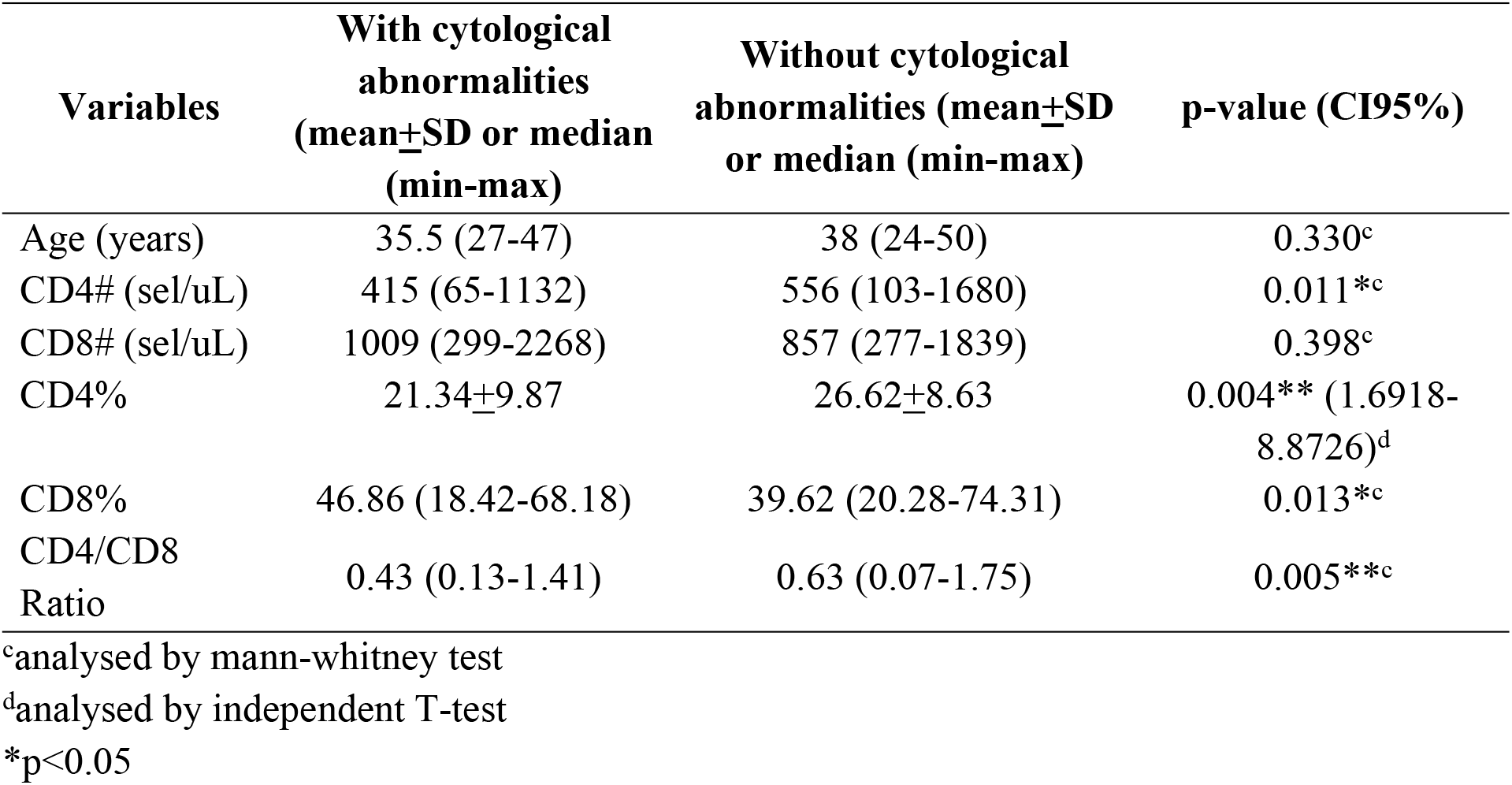
Characteristics of samples based on with and without cervical cytological abnormalities.

Since there were eight non-typeable samples, we only included 231 samples to identify the risk factors of high-risk HPV infection in women living HIV. Based on Table 3, the characteristics of samples with and without high-risk HPV infection were analyzed. It was found that marital status varied, with divorced participants making up 22.1% of those with HPV and 77.9% of those without. Married participants constituted 20.3% of the HPV-positive group and 79.7% of the HPV-negative group, while single participants represented 31.3% of those with HPV and 68.8% without. Education levels also showed differences; those with less than high school education made up 19.8% of the HPV-positive group and 80.2% of the HPV-negative group. Regarding contraception use, 25.2% of those using contraception were HPV-positive compared to 74.8% who were negative. The number of sexual partners also correlated with HPV status; 27.3% of those with more than five partners were HPV-positive.

**Table 3.**
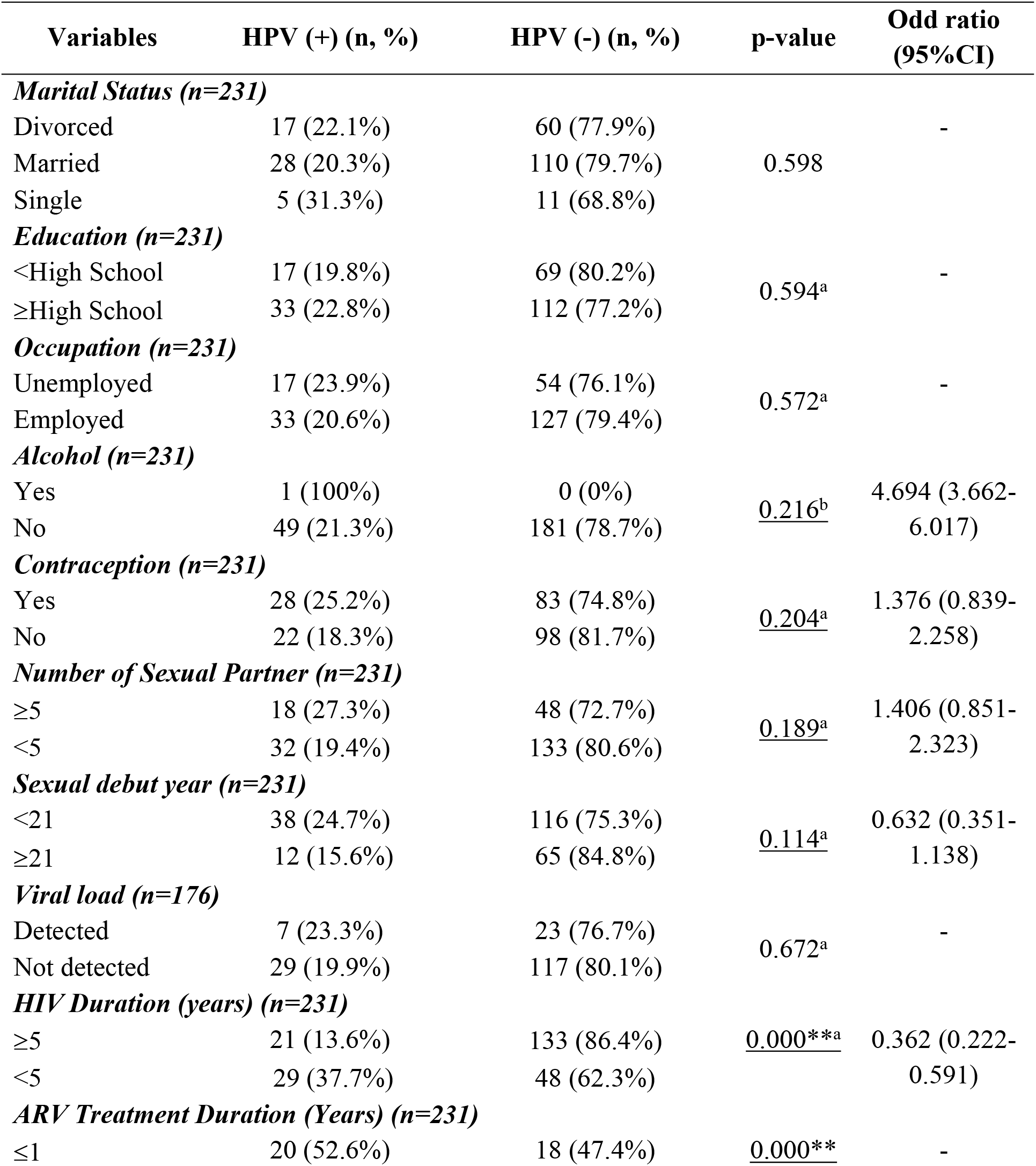
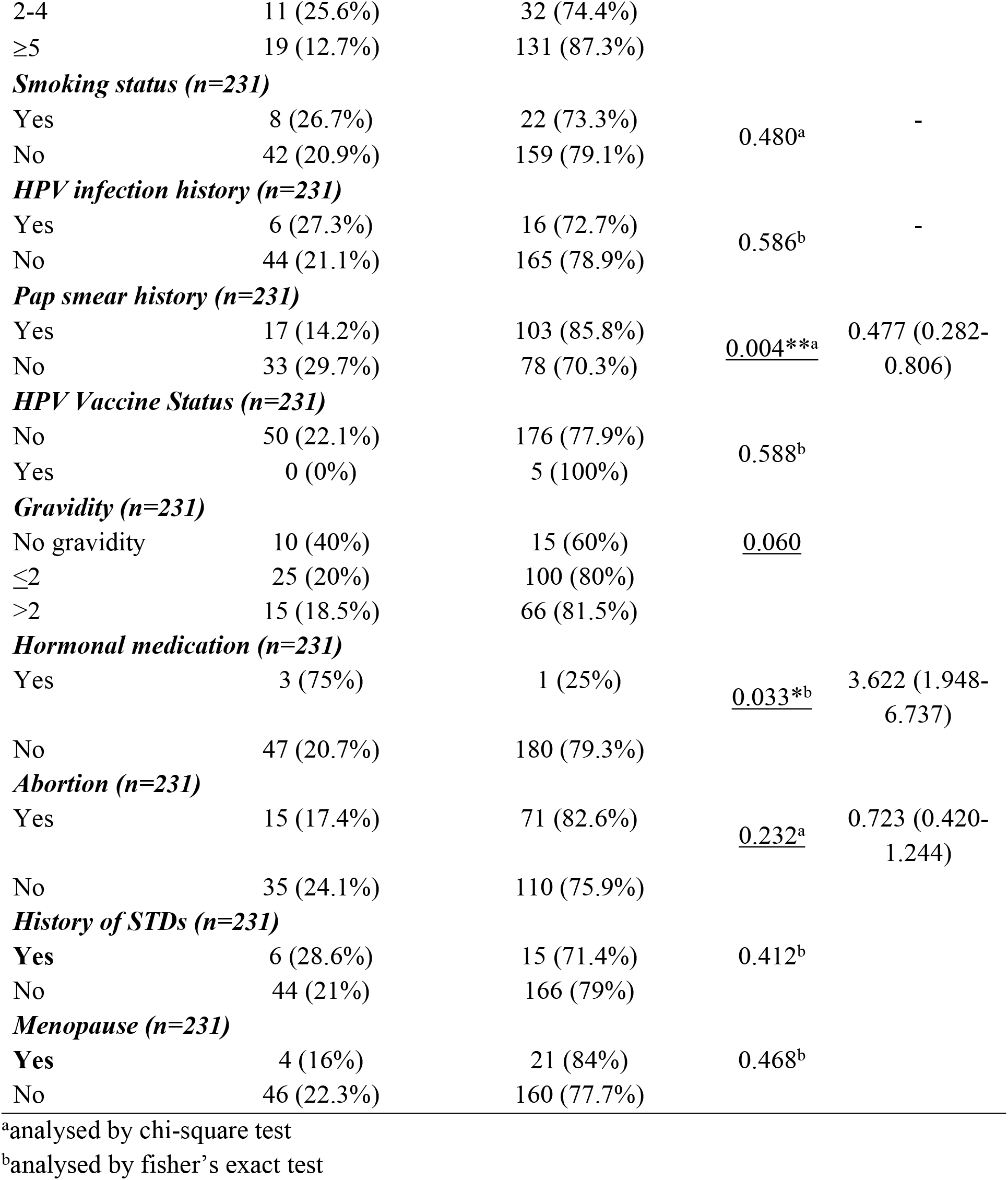
Characteristics of samples with and without high-risk HPV infection.

From the analysis presented in Table 4, key findings regarding the characteristics of samples with and without high-risk HPV infection were observed. The age of participants did not show a significant difference, with both HPV-positive and HPV-negative groups having a median age of 38 years. However, there was a significant difference in CD4 cell counts; participants with HPV had a median CD4 count of 407 cells/uL, while those without HPV had a higher median of 594 cells/uL. This suggested a lower immune status in the HPV-positive group. Additionally, the percentage of CD4 cells was significantly lower in the HPV-positive participants, averaging 21% compared to 27.44% in the HPV-negative group. The CD8 cell counts were similar between the two groups, but the CD4/CD8 ratio was significantly lower in the HPV-positive group, with a median of 0.44 compared to 0.68 in the HPV-negative group, indicating a more pronounced immune system imbalance in those with high-risk HPV infection.

**Table 4.** Characteristics of samples with and without high-risk HPV infection.

| Variables | HPV (+) (mean±SD or median (min-max)) | HPV (-) (mean±SD or median (min-max)) | p-value (CI95%) |
| --- | --- | --- | --- |
| Age (years) | 38 (24-48) | 38 (24-50) | 0.178<sup>c</sup> |
| CD4# (sel/uL) | 407 (65-1214) | 594 (103-1680) | 0.000<sup>**c</sup> |
| CD8# (sel/uL) | 881 (299-2268) | 854 (277-1839) | 0.602<sup>c</sup> |
| CD4% | 21±10.53 | 27.44±7.86 | 0.000<sup>**</sup> (-9.6311-(-3.2436))<sup>d</sup> |
| CD8% | 45.72±12.08 | 39.16 (20.28-70.13) | 0.002<sup>**c</sup> |
| CD4/CD8 Ratio | 0.44 (0.11-1.75) | 0.68 (0.07-1.54) | 0.000<sup>**c</sup> |

<sup>c</sup>analysed by mann-whitney test
<sup>d</sup>analysed by independent T-test
\*p<0.05
\*\*p<0.01

### HPV genotyping and cytology results

In the study, Table 5 and Table 6 provided insights into Pap smear and HPV genotyping results. The Pap smear results showed that 37.7% of the samples were normal, while various abnormalities were detected: 2.5% had Atypical Squamous Cells of High Grade (ASC-H), 3.8% had Atypical Squamous Cells of Undetermined Significance (ASC-US), 1.7% had High Grade Squamous Intraepithelial Lesion (H-SIL), and 2.9% had Low Grade Squamous Intraepithelial Lesion (L-SIL). The HPV genotyping revealed that HPV 18 was the most prevalent, found in 56.2% of the samples with Cytological abnormalities, significantly higher than other types. HPV 16, 51, and 52 were also detected but with lower frequencies. The results from these tables underscored the prevalence of high-risk HPV types among women with HIV and their association with various grades of cervical abnormalities.

**Table 5.**
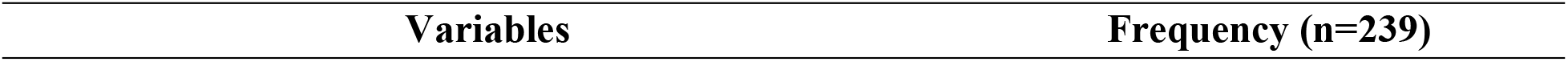
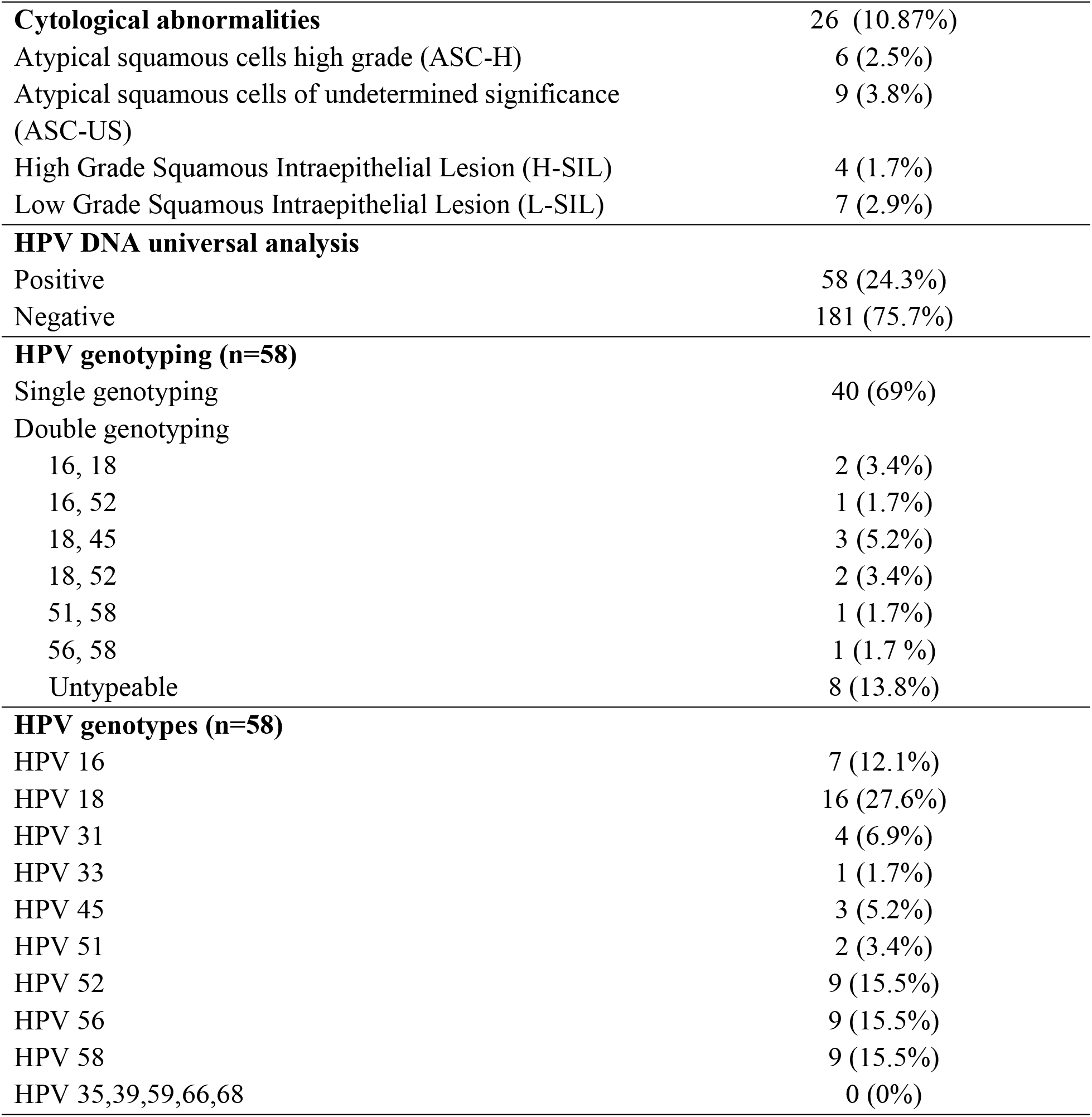
Cytological Abnormalities and HPV genotyping.

| Variables | Frequency (n=239) |
| --- | --- |
| **Cytological abnormalities** | 26 (10.87%) |
| Atypical squamous cells high grade (ASC-H) | 6 (2.5%) |
| Atypical squamous cells of undetermined significance (ASC-US) | 9 (3.8%) |
| High Grade Squamous Intraepithelial Lesion (H-SIL) | 4 (1.7%) |
| Low Grade Squamous Intraepithelial Lesion (L-SIL) | 7 (2.9%) |
| **HPV DNA universal analysis** |  |
| Positive | 58 (24.3%) |
| Negative | 181 (75.7%) |
| **HPV genotyping (n=58)** |  |
| Single genotyping | 40 (69%) |
| Double genotyping |  |
| 16, 18 | 2 (3.4%) |
| 16, 52 | 1 (1.7%) |
| 18, 45 | 3 (5.2%) |
| 18, 52 | 2 (3.4%) |
| 51, 58 | 1 (1.7%) |
| 56, 58 | 1 (1.7 %) |
| Untypeable | 8 (13.8%) |
| **HPV genotypes (n=58)** |  |
| HPV 16 | 7 (12.1%) |
| HPV 18 | 16 (27.6%) |
| HPV 31 | 4 (6.9%) |
| HPV 33 | 1 (1.7%) |
| HPV 45 | 3 (5.2%) |
| HPV 51 | 2 (3.4%) |
| HPV 52 | 9 (15.5%) |
| HPV 56 | 9 (15.5%) |
| HPV 58 | 9 (15.5%) |
| HPV 35,39,59,66,68 | 0 (0%) |

**Table 6.** Distribution of samples into cytology groups based on HPV genotypes (n=239)

| HPV genotype | With Cytological abnormalities (n, %) | Without Cytological abnormalities (n, %) | p-value |
| --- | --- | --- | --- |
| HPV 16 (+) | 3 (42.9%) | 4 (57.1%) | 0.000** |
| Other than HPV 16 | 15 (29.4%) | 36 (70.6%) |  |
| Negative for HPV | 8 (4.4%) | 173 (95.6%) |  |
| HPV 18 (+) | 9 (56.2 %) | 7 (43.8%) | 0.000** |
| Other than HPV 18 | 9 (21.4%) | 33 (78.6%) |  |
| Negative for HPV | 8 (4.4%) | 173 (95.6%) |  |
| HPV 31 (+) | 2 (50.0%) | 2 (50.0%) |  |
| Other than HPV 31 | 16 (29.6%) | 38 (70.4%) | 0.000** |
| Negative for HPV | 8 (4.4%) | 173 (95.6%) |  |
| HPV 33 (+) | 0 (0.0%) | 1 (100.0%) |  |
| Other than HPV 33 | 18 (31.6%) | 39 (68.4%) | - |
| Negative for HPV | 8 (4.4%) | 173 (95.6%) |  |
| HPV 45 (+) | 1 (33.3%) | 2 (66.7%) |  |
| Other than HPV 45 | 17 (30.9%) | 38 (69.1%) | 0.000** |
| Negative for HPV | 8 (4.4%) | 173 (95.6%) |  |
| HPV 51 (+) | 1 (50.0%) | 1 (50.0%) |  |
| Other than HPV 51 | 17 (30.4%) | 39 (69.6%) | 0.000** |
| Negative for HPV | 8 (4.4%) | 173 (95.6%) |  |
| HPV 52 (+) | 4 (44.4%) | 5 (55.6%) |  |
| Other than HPV 52 | 14 (28.6%) | 35 (71.4%) | 0.000** |
| Negative for HPV | 8 (4.4%) | 173 (95.6%) |  |
| HPV 58 (+) | 0 (0.0%) | 9 (100.0%) |  |
| Other than HPV 58 | 18 (36.7%) | 31 (63.3%) | - |
| Negative for HPV | 8 (4.4%) | 173 (95.6%) |  |

### Multivariate analysis of cervical cytological abnormalities’ risk factors

Table 7 provided a multivariate analysis of the risk factors for cervical cytological abnormalities in women living with HIV. The analysis revealed that HPV 18 was a significant independent risk factor, with an exponent (B) of 9.029 and a p-value of 0.007, indicating a strong association between HPV 18 infection and the presence of cervical abnormalities. Other HPV types such as HPV 16, 31, 45, 51, and 52 were also analyzed, but they did not show significant associations as their p-values were higher, suggesting less impact on the risk of cervical cytological abnormalities. Additionally, a history of Pap smear screening showed a trend towards reducing the risk of cervical abnormalities, although it was not statistically significant with a p-value of 0.083. The analysis underscored the critical role of HPV 18 in contributing to cervical cytological abnormalities among women with HIV and highlighted the potential protective effect of regular Pap smear screenings.

**Table 7.** The effects of all the potential risk factors on the risk of cervical cytological abnormalities in women with HIV in this study (n= 239)

| Covariates | Exponent (B) | P-value | 95% CI |
| --- | --- | --- | --- |
| HPV DNA | 2.800 | 0.177 | 0.629 – 12.45 |
| HPV 16 | 1.99 | 0.492 | 0.280 – 14.155 |
| HPV 18 | 9.029 | **0.007\*\*** | **1.835 – 44.425** |
| HPV 31 | 3.908 | 0.300 | 0.298 – 51.317 |
| HPV 45 | 0.617 | 0.745 | 0.034 – 11.327 |
| HPV 51 | 8.295 | 0.200 | 0.327 – 210.262 |
| HPV 52 | 3.277 | 0.207 | 0.519 – 20.680 |
| Papsmear history | 0.374 | 0.083 | 0.123 – 1.138 |
| ARV treatment duration ($\leq 1$ year) | 0.388 | 0.212 | 0.088 – 1.718 |
| Contraception history | 2.710 | 0.065 | 0.940 – 7.809 |
| Occupation (unemployed) | 1.845 | 0.250 | 0.650 – 5.235 |
| CD4:CD8 ratio | 0.378 | 0.234 | 0.076 – 1.876 |

### Multivariate analysis of high-risk HPV infection risk factors

Table 8 presented a multivariate analysis of the risk factors for high-risk HPV infection in women living with HIV. The findings indicated that a history of contraception use was marginally significant as a risk factor, with an exponent (B) of 2.189 and a p-value of 0.053, suggesting a potential association with increased risk of HPV infection. The number of sexual partners and the age at sexual debut did not show significant associations with HPV infection, as indicated by their p-values. Notably, a history of Pap smear screening was significantly associated with a reduced risk of high-risk HPV infection, with an exponent (B) of 0.358 and a p-value of 0.013, highlighting the protective effect of regular screening. The duration of ARV treatment and other factors like gravidity, abortion, and hormonal treatment did not show significant associations with HPV risk. This analysis underscored the importance of Pap smear history in reducing the risk of HPV infection among women with HIV.

**Table 8.**
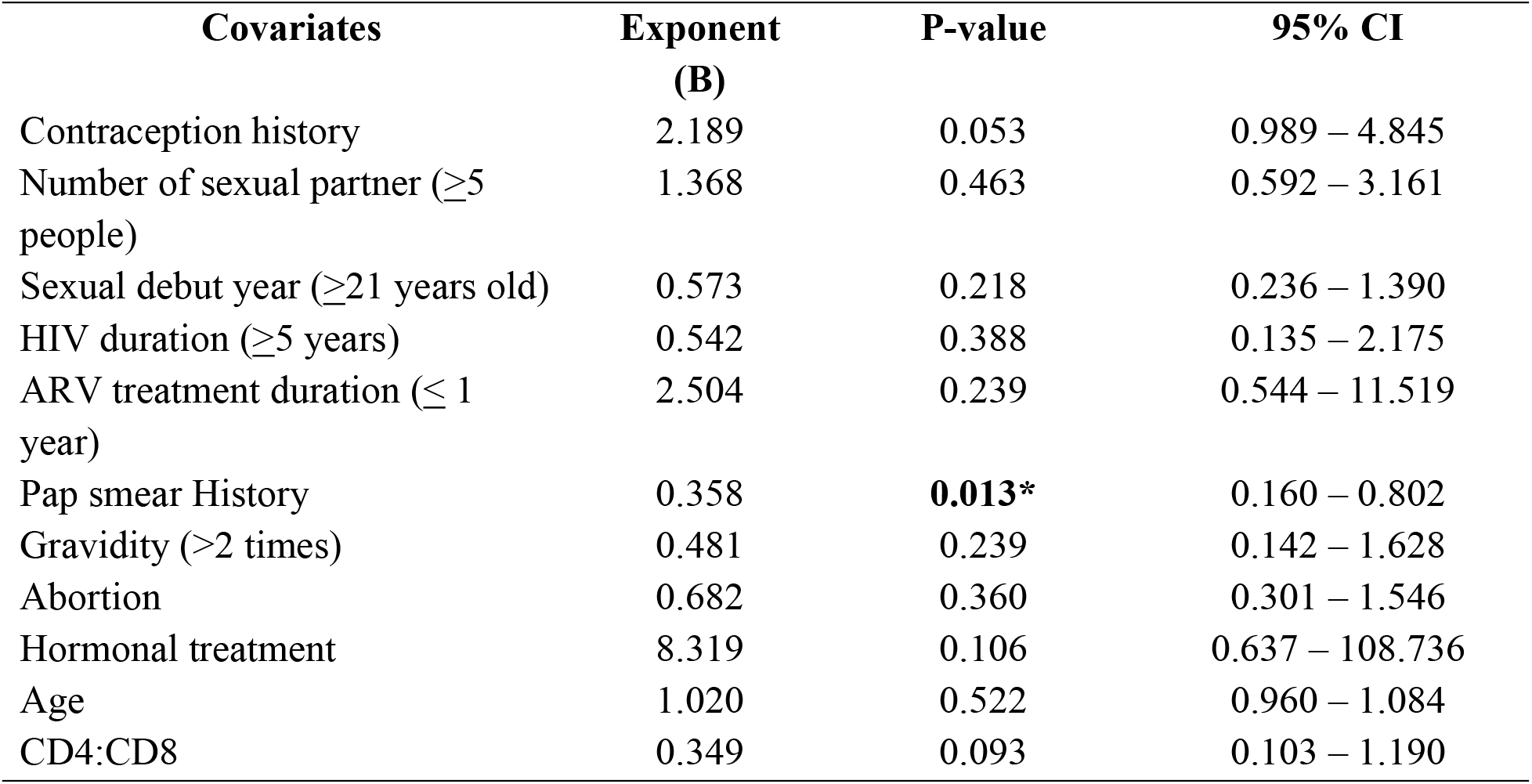
The effects of all the potential risk factors on the risk of high-risk HPV infection in women with HIV in this study (n= 231)

| **Covariates** | **Exponent**<br>**(B)** | **P-value** | **95% CI** |
| --- | --- | --- | --- |
| Contraception history | 2.189 | 0.053 | 0.989 – 4.845 |
| Number of sexual partner ($\geq 5$ people) | 1.368 | 0.463 | 0.592 – 3.161 |
| Sexual debut year ($\geq 21$ years old) | 0.573 | 0.218 | 0.236 – 1.390 |
| HIV duration ($\geq 5$ years) | 0.542 | 0.388 | 0.135 – 2.175 |
| ARV treatment duration ($\leq 1$ year) | 2.504 | 0.239 | 0.544 – 11.519 |
| Pap smear History | 0.358 | **0.013\*** | 0.160 – 0.802 |
| Gravidity ($> 2$ times) | 0.481 | 0.239 | 0.142 – 1.628 |
| Abortion | 0.682 | 0.360 | 0.301 – 1.546 |
| Hormonal treatment | 8.319 | 0.106 | 0.637 – 108.736 |
| Age | 1.020 | 0.522 | 0.960 – 1.084 |
| CD4:CD8 | 0.349 | 0.093 | 0.103 – 1.190 |

## Discussion

### HR-HPV 18 as a risk factor of cervical abnormalities in women living with HIV

Our study found that HR-HPV 18 as the only significant risk factor of cervical cytological abnormalities, potential of cervical cancer. However, a meta-analysis 2021 represents a different scenario where HPV 16 was identified as the most prevalent genotype, particularly in the regions of India, Thailand, and China.(9) Another study of meta-analysis about HPV genotypes in Kenya also showed that HPV 16 as the most common infection, with HPV 18 came second. HPV 16 was consistently identified as the most prevalent genotype in women with abnormal cytology and ICC. For instance, in women with abnormal cytology, HPV 16 was found to be present in 26% of cases, and in women with ICC, its prevalence increased to 37%.(10) This indicates a clear escalation in the prevalence of HPV 16 as the severity of cervical disease increases. Following HPV 16, HPV 18 is the second most common genotype in ICC cases, with a prevalence of 24%. This discrepancy highlights the geographical and demographic variations in HPV genotype distribution, which is a crucial factor in understanding the epidemiology of HPV and its associated risks for cervical cancer.

HPV 18, along with HPV 16, belongs to the high-risk category of HPV genotypes that are strongly associated with the development of cervical cancer. These genotypes are known for their ability to integrate into the host cell DNA, disrupting normal cell functions and leading to the progression of precancerous lesions to invasive cancer.(11) The variation in prevalence between our findings and the previous studies could be attributed to differences in study populations, methodologies, or regional public health practices such as the prevalence of vaccination and screening. This underlines the importance of localized data to inform public health strategies effectively. For regions where HPV 18 is more prevalent, targeted interventions to address this specific risk factor are crucial.(10) This includes promoting HPV vaccination that covers HPV 18, enhancing screening programs to detect and treat precancerous changes early, and educating the public and healthcare providers about the risks associated with HPV 18.

### Pap smear as an effective preventive measure of cervical cancer

The Pap smear is a critical preventive measure for cervical cancer, particularly significant for HIV-seropositive women in developing countries. As a cytological-based test, the Pap smear has been extensively utilized in developed countries for many years, demonstrating substantial success in reducing both the incidence and mortality of cervical cancer.(12)

One of the primary benefits of the Pap smear is its ability to detect precancerous changes in the cervix, known as cervical intraepithelial neoplasia (CIN), at an early stage. Early detection allows for the effective treatment of these changes before they have the chance to progress to cancer. Consequently, this early intervention significantly contributes to the reduction in mortality associated with cervical cancer, as treatments tend to be more effective at earlier stages.(5,13)

Moreover, the Pap smear is a cost-effective procedure. It does not require highly specialized equipment, which makes it a viable option for many healthcare settings, including those with limited resources typical of developing countries. The success of the Pap smear in developed countries highlights its potential effectiveness when implemented with regular screenings and proper follow-up care.(5)

Despite its benefits, the implementation of the Pap smear in developing countries faces several challenges. These include a lack of proper infrastructure, insufficient qualified personnel, and limited resources, which can impede the utilization and effectiveness of the Pap smear.(5) Nonetheless, it remains a cornerstone of cervical cancer prevention strategies. There is a pressing need to improve its accessibility and integration into healthcare systems, especially for populations at higher risk, such as HIV-seropositive women.

This study is not without limitation. The cross-sectional nature of this research did indeed restrict the ability to establish causal relationships between high-risk HPV infection and cervical cytological abnormalities. The reliance on self-reported data for certain demographic and behavioral factors might also affect the accuracy of the information collected. This study was conducted in specific region in Indonesia, that was bali, which may not account for regional variations in HPV prevalence in Indonesia; nonetheless, this study proved that in Bali, a different type of HPV was found to be more prevalent than the usual high-risk HPV found in other areas in the world.

## Conclusions

Our research found HR-HPV 18 as a significant risk factor for cervical cytological abnormalities and potentially, cervical cancer in women living with HIV, supporting the indicative measure for a routine pap-smear screening in this specified population. Along with the pap smear, vaccination for HR-HPV 18 also becomes urgently important to be readily supplied in Indonesia.

## Data Availability

Data cannot be shared publicly because of patients confidentiality. Data are available from the Institutional Data Access / Ethics Committee (contact via first author) for researchers who meet the criteria for access to confidential data.

## Authors’ contributions

We would like to thank our outstanding team who have helped us in completing this research; Study committee: TREAT Asia: Annette Sohn, Thida Chanyachukul; Kirby Institute: Kathy Petoumenos; Molecular Biology Laboratory Udayana University: Ida Bagus Nyoman Putra Dwija; Pathology Anatomy Faculty of Medicine Udayana University: Nyoman Winarti; Prof Ngoerah Hospital VCT Clinic: Komang Sutarni, Made Ratni; WM Medika Clinic - Kerti Praja Foundation Denpasar: Ni Made Vidia Pratiwi, Ni Kadek Ayu Astuti; Social workers accompanying PLWHA at VCT Prof Ngoerah Denpasar: ketut Rediten, Dedi Junaedi, Afrina Dili Astuti.

## References

1. Stelzle D, Tanaka LF, Lee KK, Ibrahim Khalil A, Baussano I, Shah ASV, et al. Estimates of the global burden of cervical cancer associated with HIV. Lancet Glob Health. 2021 Feb 1;9(2):e161–9.

2. Jin ZY, Liu X, DIng YY, Zhang ZF, He N. Cancer risk factors among people living with HIV/AIDS in China: A systematic review and meta-analysis. Sci Rep. 2017 Dec 1;7(1).

3. Rohner E, Bütikofer L, Schmidlin K, Sengayi M, Maskew M, Giddy J, et al. Cervical cancer risk in women living with HIV across four continents: A multicohort study. Int J Cancer. 2020 Feb 1;146(3):601–9.

4. WHO. Tackling NCDs: ‘best buys’ and other recommended interventions for the prevention and control of noncommunicable diseases; updated (2017) appendix 3 of the Global Action Plan for prevention and control of noncommunicable diseases 2013–2020. 2017;

5. Robbers GML, Bennett LR, Spagnoletti BRM, Wilopo SA. Facilitators and barriers for the delivery and uptake of cervical cancer screening in Indonesia: a scoping review. Vol. 14, Global Health Action. Taylor and Francis Ltd.; 2021.

6. Kementeian Keesehatan RI Indonesia. Kementerian Kesehatan RI. Profil Kesehatan Indonesia 2016 Kementrian Kesehatan Indonesia [Internet]. 2017.

7. Ogu CO, Achukwu PU, Nkwo PO. Prevalence and risk factors of Cervical Cytological abnormalities among human immunodeficiency virus sero-positive females on highly active antiretroviral therapy in Enugu, Southeastern, Nigeria. Asian Pacific Journal of Cancer Prevention. 2019;20(10):2987–94.

8. WHO. WHO guideline for screening and treatment of cervical pre-cancer lesions for cervical cancer prevention. 2021.

9. Verrier F, Le Coeur S, Delory T. Cervical human papillomavirus infection (Hpv) and high oncogenic risk genotypes among women living with hiv in asia: A meta-analysis. Vol. 10, Journal of Clinical Medicine. MDPI; 2021.

10. Menon S, Wusiman A, Boily MC, Kariisa M, Mabeya H, Luchters S, et al. Epidemiology of HPV genotypes among HIV positive women in Kenya: A systematic review and meta-analysis. PLoS One. 2016 Oct 1;11(10).

11. Freitas BC, Suehiro TT, Consolaro MEL, Silva VRS. HPV infection and cervical abnormalities in HIV positive women in different regions of Brazil, a middle-income country. Asian Pacific Journal of Cancer Prevention. 2016;16(18):8085–91.

12. Mapanga W, Girdler-Brown B, Feresu SA, Chipato T, Singh E. Prevention of cervical cancer in HIV-seropositive women from developing countries through cervical cancer screening: A systematic review. Vol. 7, Systematic Reviews. BioMed Central Ltd.; 2018.

13. Asangbeh-Kerman SL, Davidović M, Taghavi K, Kachingwe J, Rammipi KM, Muzingwani L, et al. Cervical cancer prevention in countries with the highest HIV prevalence: a review of policies. BMC Public Health. 2022 Dec 1;22(1).

